# A Graph Based Deep Learning Framework for Predicting Spatio-Temporal Vaccine Hesitancy

**DOI:** 10.1101/2023.10.24.23297488

**Authors:** Sifat Afroj Moon, Rituparna Datta, Tanvir Ferdousi, Hannah Baek, Abhijin Adiga, Achla Marathe, Anil Vullikanti

## Abstract

Predicting vaccine hesitancy at a fine spatial level assists local policymakers in taking timely action. Vaccine hesitancy is a heterogeneous phenomenon that has a spatial and temporal aspect. This paper proposes a deep learning framework that combines graph neural networks (GNNs) with sequence module to forecast vaccine hesitancy at a higher spatial resolution. This integrated framework only uses population demographic data with historical vaccine hesitancy data. The GNN learns the spatial cross-regional demographic signals, and the sequence module catches the temporal dynamics by leveraging historical data. We formulate the problem on a weighted graph, where nodes are zip codes and edges are generated using three distinct mechanisms: 1) adjacent graph - if two zip codes have a shared boundary, they will form an edge between them; 2) distance-based graph - every pair of zip codes are connected with an edge having a weight that is a function of centroid distances, and 3) mobility graph - edges represent the number of contacts between any two zip codes, where the contacts are derived from an activity-based social contact network. Our framework effectively predicts the spatio-temporal dynamics of vaccine hesitancy at the zip-code level when the mobility network is used to formulate the graph. Experiments on the real-world vaccine hesitancy data from the All-Payer Claims Database (APCD) show that our framework can outperform a range of baselines.

## 1 Introduction

Highly contagious diseases, such as measles, are regarded as vaccine-preventable (VPD) because of the availability of the Measles, Mumps, and Rubella (MMR) vaccine, which has a very high efficacy rate. Measles is preventable using high rates of immunization. The MMR vaccine is required by public schools in most parts of the world, including the US, and measles was declared as “eliminated” from the US in 2000 (Centers for Disease Control 2020). Unfortunately, immunization rates are declining for many childhood vaccines, and outbreaks of measles and other VPDs have been occurring regularly in recent years across the world. For instance, there was a large outbreak in New York in 2019, which caused over 900 cases (Patel et al. 2019). In 2021, Nigeria had over 10,000 cases (Sato et al. 2022), and there were 128,000 deaths due to measles worldwide (Organization 2023). The risk of measles, and other vaccine-preventable diseases, has significantly been exacerbated due to the COVID-19 pandemic (Thakur et al. 2022).

There are a number of reasons behind the drop in immunization rates, and hesitancy is the leading among them. Even before the pandemic, though the MMR vaccine coverage was quite high (∼95%) for kindergarten children nationally (Seither et al. 2023) (which is a high enough rate to reach herd immunity), it was not evenly spread geographically, and there were significant pockets of undervaccination (Lieu et al. 2015; Cadena et al. 2019a; Gahr et al. 2014; Gastañaduy et al. 2016). During the pandemic, significant drops in routine immunizations have been reported (Seither et al. 2022; Causey et al. 2021; Iacobucci 2022) In 2020 and 2021, over 27 and 25 million children were estimated to have missed their first dose of the measles vaccine, respectively (Causey et al. 2021; Guglielmi 2022). Measles is now viewed as an imminent global threat (Iacobucci 2022).

Vaccine hesitancy is a growing concern in public health (McGregor and Goldman 2021), and predicting vaccine hesitancy at the higher spatial resolution is considered a fundamental problem, as heterogeneous vaccine coverage significantly increases the risk of outbreaks (Masters et al. 2020; Moon, Marathe, and Vullikanti 2023). One of the significant challenges in understanding the extent of hesitancy and how it is spreading is the limited availability of surveillance data on declining immunization rates, especially at finer spatial resolutions. Surveys on immunization rates are often available at a coarse resolution (e.g., a state) (Seither et al. 2022; Causey et al. 2021), which does not help identify specific under-immunized regions for intervention. Many states provide School Immunization Survey (SIS) reports (of Health 2021), which provide immunization rates for schools. However, they only consider specific age groups (e.g., 4-6 years) and miss a lot of children (e.g., those who are home-schooled).

*The focus of our paper is to develop methods to predict vaccine hesitancy at the zip code level*; we refer to this as the VaccHesitancy problem. In this work, we investigate vaccine hesitancy among kids aged between 0-6 years. Prior works have used this information as a reflection of parental vaccine intention (Müller, Tellier, and Kurschilgen 2022). Kids in this age range are expected to receive a set of mandatory vaccines including MMR (Measles, Mumps, and Rubella), HepB (Hepatitis B), and DTaP (Diphtheria, Tetanus, and Pertussis). A novel aspect of our work is the use of an extensive insurance claims dataset for Virginia that includes all insurance claims for over 5 million individuals over a five years period.

Analyzing vaccine hesitancy has been an active area of research. Many works have focused on understanding hesitancy and identifying the responsible factors using social media data (Nguyen et al. 2022). However, these data contain notable biases from demographic variations in platform preferences and the information individuals opt to share. On the other hand, some recent data-driven approaches use detailed individual-level data, making it harder to generalize. We discuss the relevant works on vaccine hesitancy modeling in Section 2.

Our main contributions are as follows:

- We develop a novel approach, VH-GNN, for the VaccHesitancy problem by combining a GNN and a recurrent neural network (RNN) using demographic data and historical hesitancy data, along with detailed population mixing data in the state. The GNN captures the spatial aspect of vaccine hesitancy by learning the impact of neighboring zip codes with respect to population-level mixing. The RNN learns temporal dynamics by leveraging historical hesitancy data. In the rest of the paper, we refer to the combined framework as VH-GNN; Vaccine Hesitancy predicted Graph Neural Network. Figure 1 shows the VH-GNN architecture.
- We train and evaluate the VH-GNN using the large-scale insurance claims data set for the state of Virginia, mentioned earlier, and show that our model outperforms a number of baselines, leading to a substantial reduction in prediction errors ranging from 18.40% to 43.4%.
- Through an ablation study, we demonstrate the effectiveness of the combined framework in improving model performance. In particular, we find that spatial structure from the detailed population-level mixing is very significant in forecasting vaccine hesitancy. We explore other kinds of connectivity and spatial structure too, but do not find them to be as predictive.
- While the performance of VH-GNN is generally high, we find there are some zip codes (denoted by set *V*_*L*_) where the prediction error is high. We identify several features which characterize the zip codes in *V*_*L*_ such as, Kids population, vaccine hesitancy percentage, medicaid insurance percentage, hispanic population percentage.
- In order to understand the structure of the solution from VH-GNN and the true hesitancy level datasets, we use the Moran’s-I and isolation indices, which are metrics for quantifying spatial clustering. We find that Moran’s-I is high and the isolation index is low, indicating a similar clustering structure between the solution predicted by VH-GNN and the actual claims dataset.

## 2 Related Work

Research on vaccine hesitancy prediction can be divided into two major categories: data-driven studies and model-based studies. Recent data-driven vaccine hesitancy studies explore different machine learning models, such as neural networks, random forest, logistic regression, recursive partitioning, and support vector machines, to find local vaccine hesitancy hotspots or to predict individual decisions (Carrieri, Lagravinese, and Resce 2021; Chandir et al. 2018; Bell et al. 2019). These studies do not consider the spatial aspect of vaccine hesitancy. However, vaccine refusal has a spatial clustering nature, and the immunization status of kids shows correlations in the same neighborhood, schools, or jurisdictions (Atwell et al. 2013; Lieu et al. 2015; Nsoesie et al. 2013). In addition, these studies have used detailed socio-economic data, including siblings’ vaccination history and private medical history, which are inaccessible in many cases due to privacy concerns. In this work, we only use features developed at the zip code level, such as population size, gender, race, and insurance type.

**Figure 1:**
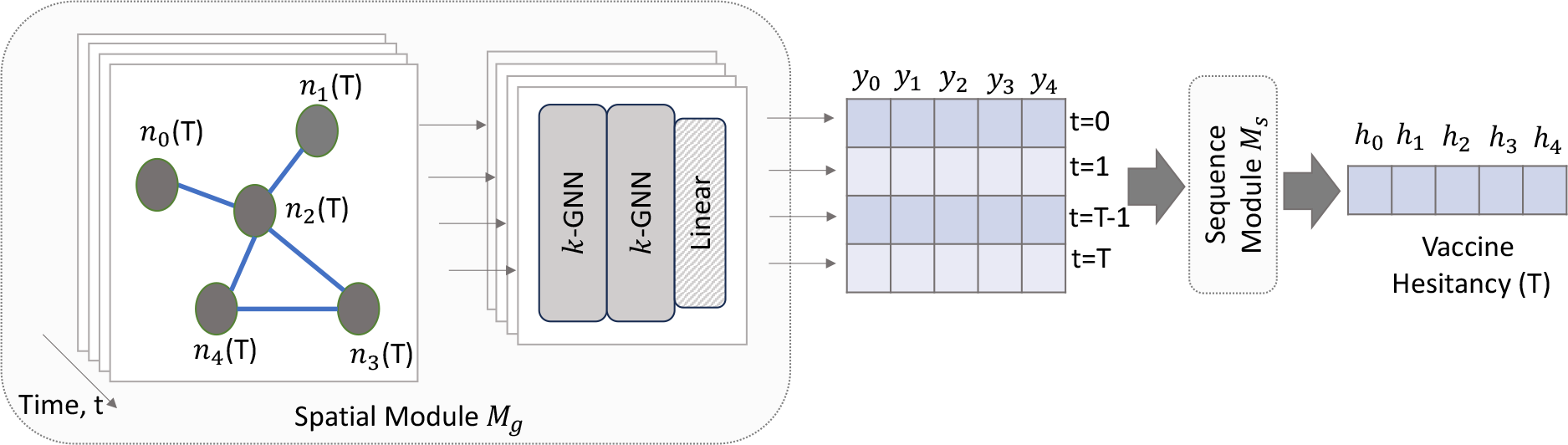
Architecture of the spatio-temporal graph-based node-level regression learning for the prediction of vaccine hesitancy.

Spatial modeling of COVID-19 vaccine hesitancy explains variations in vaccination rates using multiscale geographically weighted regression (Mollalo and Tatar 2021). A recent spatial mathematical model of opinion dynamics with reinforcement explains the occurrence of vaccine hesitancy. Mathematical models often do not consider heterogeneous social connectivity.

To address the spatial neighbor impact and temporal dynamics of vaccine hesitancy, we use a graph-based deep learning framework. It combines Graph Neural Network (GNN) with a sequence module for a node-level prediction task. GNN is a deep-learning tool specialized to handle graph data (Xu et al. 2018). GNN is widely used in different domains for graph-related prediction tasks, such as node classification, link prediction, and graph classification. GNNs have demonstrated good prediction capabilities for spatial data, such as house price estimation, understanding election results (Klemmer, Safir, and Neill 2023), and weather forecasting. In this graph-based research, nodes represent zip codes, and edges represent the connectivity among zip code pairs.

Patient refusal or vaccine hesitancy sentiment is changing with time. To learn the temporal aspect of the vaccine hesitancy levels for a location, we propose a sequence module to handle time series data for each zip code. Our framework uses a recurrent neural network structure with Gated recurrent units (GRUs) (Cho et al. 2014) as the sequence module. Spatio-temporal graph learning has been used in recent times to forecast traffic flow (Li and Zhu 2021), disease prevalence (Wang et al. 2022), etc. Prior works have used different combinations of GNN, RNN, or Convolutional Neural Networks (CNN) to perform spatio-temporal forecasting tasks (Yu, Yin, and Zhu 2017; Hu et al. 2022). Traffic forecasting is an example of spatio-temporal modeling. Yu et al. show the potential of graph-based learning frameworks for timely and accurate traffic forecasts with comparisons between CNN and RNN. We extended these concepts to predict vaccine hesitancy for a zip code in the next time step. For spatio-temporal learning, we use a static network, where nodes are zip codes, and edges are connections among pairs of zip codes. Nodes have time-varying features as attributes. Using demographic and historical vaccine hesitancy data, we forecast the vaccine hesitancy percentage of a node or a zip code.

## 3 Methods

### 3.1 Preliminary

Our goal is to learn vaccine hesitancy, measured by the percentage of patients refusing to take the vaccine within a zip code. As inputs, we use historical vaccine hesitancy data and demographic features of a zip code (which includes age, gender, race, ethnicity, and insurance type), and spatial zip code level connectivity information of several types.

Let *G*(*V, E, A*) denote a graph, where *V* denotes the set of *N* zip codes. We consider three types of connectivity information in defining the edges: spatial adjacency, the distance between zip codes, and mixing through an activity-based network (described below in Section 3.3). Let *A*^*N×N*^ denote the weighted adjacency matrix. An entry of *A, a*(*i, j*), represents the connection from node *i* to *j*. A node *i* is associated with a feature vector *x*_*i*_(*t*), which is time-dynamic. here, *X*^*T ×N×k*^ is the feature matrix at time *t*, and *k* is the number of features.

#### The VaccHesitancy problem

Let *h*_*i*_(*t*) denote the fraction of patients indicating hesitancy in zip code *i* at time *t* (which will be specified later in Section 4); let **h**(*t*) denote the vector of hesitancy levels. The VaccHesitancy problem we involve learning **h**(*T*) using historic hesitancy levels **h**(*t*^′^), demographic characteristics of zip codes in *V*, and the connectivity network *G*.

### 3.2 Framework

An analysis of the hesitancy levels **h**(*t*) over time from the dataset reveals spatial heterogeneity and correlations (presented in the Appendix), which motivates our GNN approach based on spatial structure. Our VH-GNN framework (Algorithm 1) has two major modules: 1) a spatial module, and 2) a sequence module (Figure 1). The spatial module consists of graph forming and graph-based spatial dependency learning, described in Section 3.3.

### 3.3 Spatial Module

#### Graph Architecture

The graph *G* is a static graph without self-loops. We propose three intuitive node connectivity mechanisms to form three variants of *G*.

- Adjacent graph *G*_*a*_: If node *i* and node *j* share a geographic boundary, they will form an edge in the network *G*. The weights of edges in the adjacent graph *G*_*a*_ are always 1.
- Distance-based graph *G*_*d*_: We have a fully connected graph, *G*_*d*_. A connection between two nodes *i* and *j* has a weight equal to the inverse of the distance between centroids of zip codes *i* and *j*.
- Mobility graph *G*_*m*_: This graph represents population movement. We form *G*_*m*_ from an activity-based detailed population-level social contact network *G*_*p*_ (Eubank et al. 2004; Barrett et al. 2009; Cadena et al. 2019b). The nodes in *G*_*p*_ represent individual people, and each node is associated with a location. We aggregate this network at the zip code level to get *G*_*m*_. If there is any connection from the population of zip code *i* to zip code *j*, we form an edge in the graph *G*_*m*_. This edge has an associated weight equal to the total number of connections from zip code *i* to *j* in the graph *G*_*p*_.

All edge weights are normalized using min-max normalization.

#### Graph-based Spatial Dependency Learning

Learning the spatial distribution of vaccine hesitancy with demographic characteristics is a major task. We leverage graph neural network (GNN) (Hamilton, Ying, and Leskovec 2017) to learn spatial dependency for each time step through message passing. In this work, node features change with time, but the graph connectivity is static. For each time step, we have one GNN module, which consists of stacking multiple k-GNN layers (Morris et al. 2019) and a linear layer to perform node-level vaccine hesitancy prediction. We model it as a regression task. The k-GNN is a generalization of graph neural networks based on the k-dimensional Weisfeiler-Leman algorithm (k-WL). This variant of GNN performs message passing directly between subgraphs instead of individual nodes. The *l*^*th*^ layer of the first-order GNN is

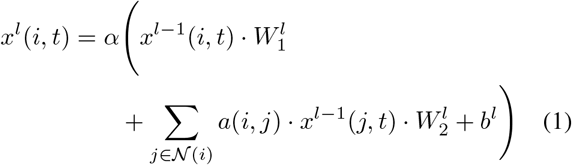

Here, *l >* 0, *α* represents an activation function (e.g., sigmoid or ReLU). 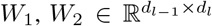 are weight matrices parametrizing GNN layer *l*, 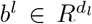 is the parameters of the l-th layer, *d*_*l*_ is the dimension, and 𝒩(*i*) is the neighborhood of *i*. The output of the graph embedding module is an intermediate solution 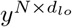 for each time point *t. lo* is the final layer of the graph embedding module. For *T* time steps, we merge the *y* matrix to form 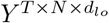.

### 3.4 Sequence Module

To learn the temporal aspect of the vaccine hesitancy for a node *i*, we use a sequence module. The input of this module is matrix *Y*, which will predict vaccine hesitancy at time *T*. This module can be built using any model that can learn sequences; popular choices are moving average, ARIMA, and recurrent neural networks. Recurrent neural networks (RNN) are well-known for predicting sequence data. In this work, we leverage a variant of RNN known as Gated Recurrent Units (GRU) (Zhao et al. 2019). We also experiment with LSTM (Long Short-Term Memory) and moving average. The fundamental concepts underlying LSTM and GRU models are quite similar. Both employ gated mechanisms to retain extensive long-term information, making them equally proficient for diverse tasks. We find that GRU performs better in the VH-GNN framework. It trains faster with fewer parameters compared to the LSTM variant. Hence, it has better potential to learn from large multidimensional datasets.

### 3.5 Optimization

This framework optimizes two modules separately with optimizers *opt*_1_ and *opt*_2_. We use two loss functions to reduce the error between predicted vaccine hesitancy *H*(*T*) and the true vaccine hesitancy *Ĥ*(*T*). The first loss function *loss*_1_ minimizes the error for the graph learning module at each time step *t* = 0, 1, .., *T*, and the second loss function *loss*_2_ reduces errors for the sequence learning module. We use the mean absolute error (*MAE*) metric to learn the model parameters.

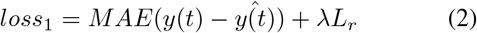

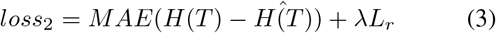

*loss*_1_ takes into account all time steps, and *loss*_2_ takes only the final time step. In this research problem, for any time step 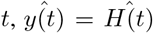, as the graph learning module is predicting vaccine hesitancy for that time step from *x*_*i*_(*t*). Here, *λ* is a hyper-parameter, and *L*_*r*_ is the *L*2 regularization term to prevent over-fitting. We optimize two modules separately as we do not want to influence one module’s parameters due to the other module’s performance. AAlgorithm 1 details the steps taken.

## 4 Experiments and Results

### 4.1 Data

We use five years (2016-2020) of the All-Payer Claims Database (APCD) to find the patient refusal levels for each zip code in Virginia.

#### Algorithm 1: VH-GNN for Spatio-Temporal Vaccine Hesitancy Learning

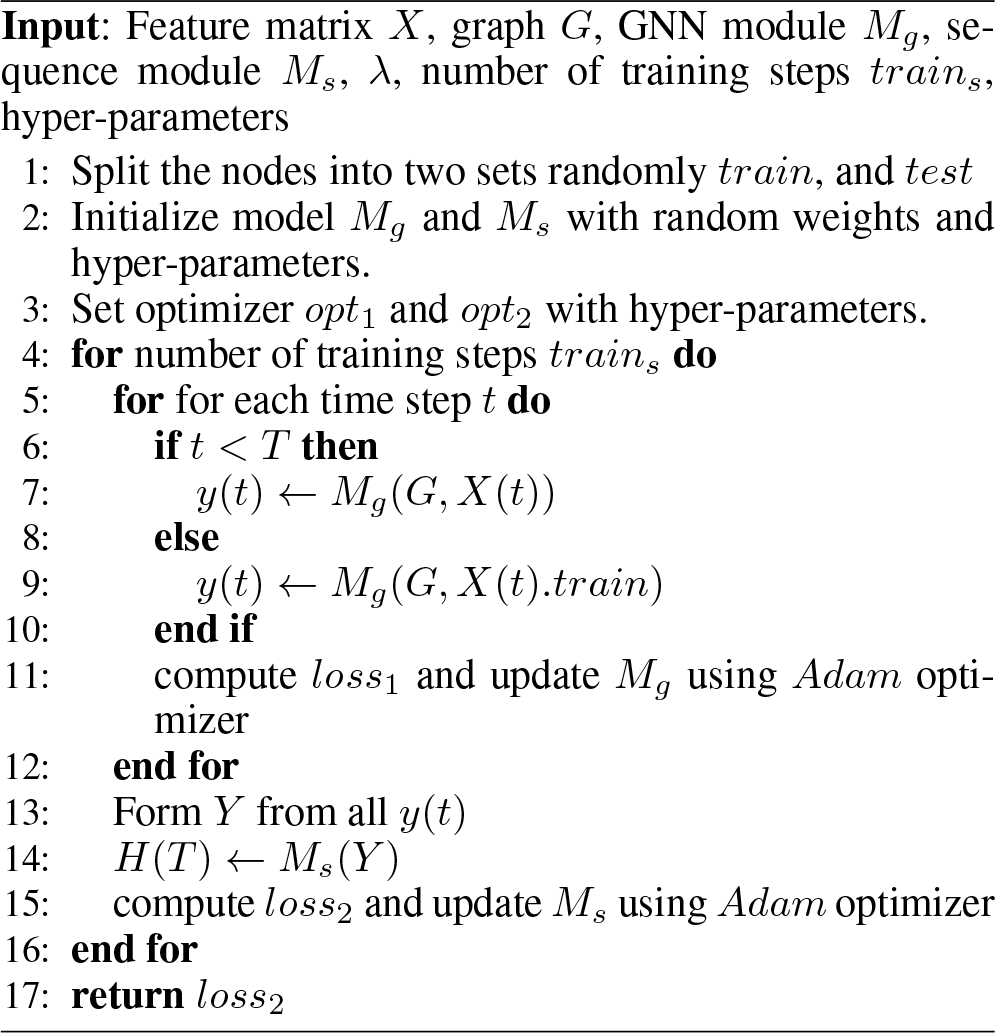

The data is obtained from VHI (Virginia Health Information). It contains information on paid medical and pharmacy claims for roughly 5 million Virginia residents with commercial, Medicaid, and Medicare coverage across all types of healthcare services. Among other things, it provides information on immunization rates over time, by spatial regions, and by demographics.

International Classification of Disease ICD-10-CM code Z28 is used to filter Patient refusal from medical data. Z28 means, “Immunization not carried out and underimmunization status” (ICD10data 2023). We also analyze the immunization rates as provided in the Virginia Department of Health School Immunization Survey (VDH SIS) reports. We find that vaccine hesitancy is changing in Virginia (Table 1). However, we only use APCD for VaccHesitancy, as our targeted population is 0-6 years old, and VDH-SIS does not contain this information. In this work, we use six months as the time unit. We find that monthly data is sparse at the zip code level. We explain more about the data in Appendix.

**Table 1:** Yearly patient refusal percentage in Virginia in the APCD data and in the VDH SIS reports.

| Vaccine Hesitancy | Year |  |  |  |  | Source |
| --- | --- | --- | --- | --- | --- | --- |
|  | 2016 | 2017 | 2018 | 2019 | 2020 |  |
| Patient Refusal | 1.55% | 3.31% | 4.01% | 4.08% | 3.83% | APCD |
| State Public School Unimmunized or not Adequately Immunized | 3.6% | 3.9% | 3.7% | 3.6% | 11.9% | VDH SIS |
| State Private School Unimmunized or not Adequately Immunized | 5.8% | 6.7% | 6.7% | 4.8% | 7.6% | VDH SIS |

In this paper, ‘zip code’ refers to a ZIP Code Tabulation Area (ZCTA). A ZCTA corresponds to a geographical representation of a service area for a United States Postal Service (USPS) ZIP Code. This delineation is made publicly available by the US Census Bureau (U.S. Census Bureau 2021). We use 615 zip codes of Virginia (*N* = 615) out of 1241. Among them, only about 52% zip codes have a population size of more than 1000. We discard zip codes that do not have any entry for kids (aged 0-6) in the APCD data or have a very small population size.

### 4.2 Data Preprocessing

From the APCD data, we filter all patients’ entries of children aged six or below. Then we prepare a data set for each time *t* for *N* nodes, which keeps a record of the number of unique kids, the number of unique kids in different genders, the number of unique kids in different races, the number of unique kids in two different medical insurance types (commercial and Medicaid), and the percentage of kids who refuse to take any vaccine at least once. We use “medical insurance type” as a proxy for the income level. We assume that patient refusal at this age represents parental vaccine intention.

At each time step *t*, the last column of the data set is the target value; vaccine hesitancy *Ĥ*(*t*), other columns are features of nodes. A node *i* has ten features at a time step *t*, including male population, female population, Asian population, Black population, White population, and Hispanic population. We use min-max normalization to normalize all columns for each time step *t* as we update our *M*_*s*_ module. Then, we find the principal components of the features by using Principal Component Analysis (PCA).

### 4.3 Experimental Setup

This study uses the GNN version of Morris et al. (Morris et al. 2019). We also explored other strategies, such as Graph Convolutional Network (GCN) (Kipf and Welling 2016), which can handle weighted static graphs. However, we find that the GNN of Morris et al. performs better in predicting vaccine hesitancy.

We manually adjust the hyper-parameters of the framework, such as the learning rate, the regularization term, the training epoch, and the number of hidden units. We use a learning rate of 0.0005, a training epoch of more than 10,000, and a learning rate of 10^−4^. We find that using more than two GNN layers overfits the training data while using less than two introduces bias in the system. For module *M*_*g*_, we experiment on hidden units [64,128,256,512]. For module *M*_*s*_, we experiment on hidden units [8,16,32,64]. The setup for *M*_*g*_ with 128 hidden units and *M*_*s*_ with 16 hidden units performs better for the VH-GNN. For all GNN layers and GRU layers, we implement 50% dropout to avoid overfitting.

We use Python 3.8 to implement the framework. We utilize the open-source deep-learning framework PyTorch version 2.0.0 and NVIDIA CUDA 11.4.2 in a Simple Linux Utility for Resource Management (SLURM) system.

### 4.4 Evaluation Metrics

We use the following two metrics to evaluate the performance of the VH-GNN framework in predicting spatiotemporal vaccine hesitancy levels for the test data set.

- Root Mean Squared Error (RMSE):

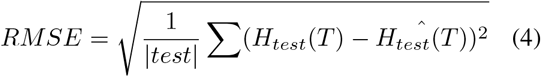
- Mean Absolute Error (MAE):

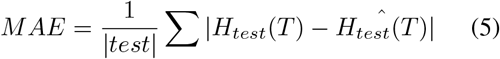

*MAE* is only used during model training. For the test data set, *RMSE* and *MAE* metrics are utilized to gauge the model’s predictive performance. Smaller values of *RMSE* and *MAE* indicate better prediction accuracy.

### 4.5 Baseline Methods

The performance of our combined graph framework is compared with the following baseline methods:

- **Linear Regression with Neighbors (LRN):** We first evaluate our model’s performance against the Linear Regression approach, where this baseline method is used to predict the dependent variable *H*(*T*) using input features. Linear regression fits a linear model to minimize the residual sum of squared differences between true and predicted values using linear approximation. To make a fair comparison, we provide an extra feature, neighbor’s information, for each node *i*, we calculate ∑_*j*∈𝒩 (*i*)_ *h*_*j*_(*t*)*a*(*i, j*).
- **Multi-layer Perceptron (MLP):** We also experiment with this second benchmark method, Multi-layer Perceptron (MLP), to assess our model’s effectiveness. This feedforward neural network architecture is capable of capturing complex non-linear relationships within data.
- **Graph Convolutional Network (GCN):** GCN is our third benchmark method. It uses convolutional architecture to capture both local and global patterns within graph-structured data for semi-supervised learning.
- **Graph Convolutional Network & Gated Recurrent Units (GCN-GRU):** In this fourth benchmark, we replace *M*_*s*_ module with the GCN in the VH-GNN.

Figure 2 shows the mean output performances of all the baselines compared to the VH-GNN framework for the target years 2019 and 2020. The same *train* and *test* data sets were used for all models. We always use a batch gradient process to update the model parameters. We employ model-specific hyperparameters to unlock their full potential. Results indicate that VH-GNN outperforms all baselines in both evaluation metrics. It is also evident that the VH-GNN performed better for the year 2019 compared to 2020.

**Figure 2:**
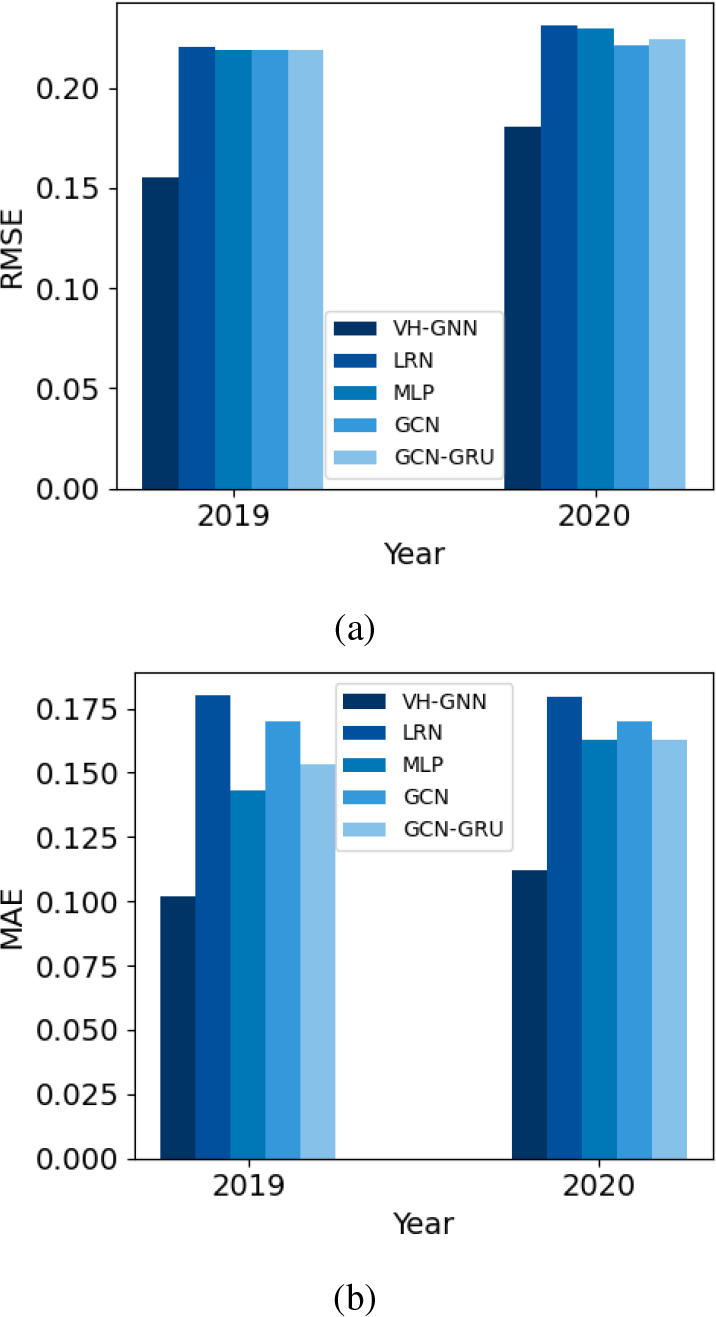
A comparative analysis between our VH-GNN and the baseline methods across 2019 and 2020. Figure 2a and 2b show that our method outperforms alternative approaches, as evident in both the RMSE and MAE metrics.

The results are also affected by the choice of node connectivity mechanisms. Table 2 shows the performance of VH-GNN using three *G* variants for 2019. The VH-GNN performs the best when mobility graph *G*_*m*_ is used. Hence, the remaining set of results in this paper for VH-GNN is produced using *G*_*m*_.

**Table 2:** Prediction performance of VH-GNN across three graphs connectivity mechanisms for the year 2019.

| Graph | RMSE | MAE |
| --- | --- | --- |
| Adjacent Graph $G_a$ | 0.2047 | 0.1487 |
| Distance-based Graph $G_d$ | 0.1816 | 0.1342 |
| Mobility Graph $G_m$ | **0.1554** | **0.1019** |

Figure 3 shows predicted values vs true values for 2019.

**Figure 3:**
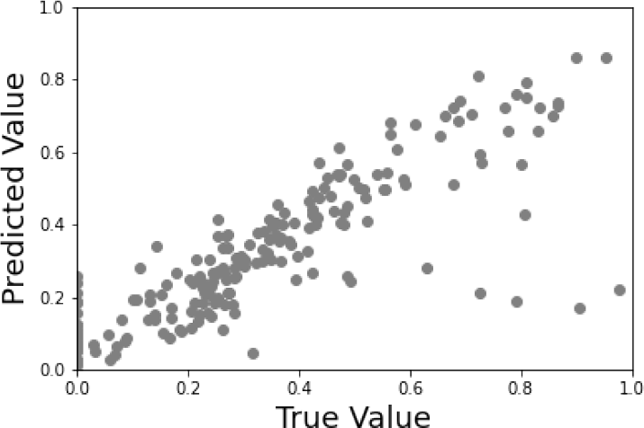
Predicted vaccine hesitancy and the true vaccine hesitancy in the *test* set for year 2019.

The *R*^2^ value is 0.61.

### 4.6 Ablation Study

We conduct an ablation study to understand the role of two modules of the VH-GNN.

- **VH-GNN w/o** *M*_*s*_ **module:** In this setup, we only train the spatial learning module *M*_*g*_ with the *loss*_1_ function and evaluate the prediction performance only using *M*_*g*_.
- **VH-GNN w/o** *M*_*g*_ **module:** In this setup, we only keep the sequence learning module, which is the GRU model. Here, we train *M*_*s*_ by using 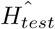 for *t* = 0 to *t* = *T* − 1 and 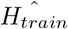 for *t* = 0 to *t* = *T*. It does not consider any graph structure.

Table 3 shows the prediction performance of the two modules. Ablation study shows the importance of spatial learning for node-level vaccine hesitancy forecasting. Although our combined framework performs better than either of these configurations, the VH-GNN w/o *M*_*s*_ outperforms VH-GNN w/o *M*_*g*_. This indicates the importance of the spatial component in explaining vaccine hesitancy.

**Table 3:** Ablation study on two setups for the year 2019, 1) VH-GNN w/o *M*_*s*_ module, and 2) VH-GNN w/o *M*_*g*_ module.

| Ablation Study Setting | RMSE | MAE |
| --- | --- | --- |
| VH-GNN w/o $M_s$ module | 0.2054 | 0.1434 |
| VH-GNN w/o $M_g$ module | 0.4557 | 0.3820 |

### 4.7 Performance Analysis at the Node Level

The properties of nodes were investigated where VH-GNN performed poorly. For a comparative analysis, the nodes are divided into two sets, one where VH-GNN performs poorly and the other where it performs well. The test nodes are sorted according to the absolute error between predicted and true vaccine hesitancy values, and the nodes were divided into two sets:

- **Nodes with Large Error**, *V*_*L*_: Top 25% nodes, nodes with large error, where VH-GNN did not perform well.
- **Nodes with Small Error**, *V*_*S*_: Rest of the test nodes, where VH-GNN performs well.

We investigated features of two sets: *V*_*L*_ and *V*_*S*_, to see why the VH-GNN framework does not predict well. We find that population sizes, vaccine hesitancy percentages, population percentages with Medicaid insurance, and Hispanic populations differ between these two sets. Table 4 reports the average of these features for *V*_*L*_ and *V*_*S*_ sets. Table 4 shows that the kids’ population sizes are significantly different across *V*_*L*_ and *V*_*S*_. The VH-GNN is prone to have large predictive errors for nodes that have a small population with a high vaccine hesitancy level. Further investigation in Figure 4 shows that VH-GNN also performs well when the vaccine hesitancy percentage is high.

**Table 4:**
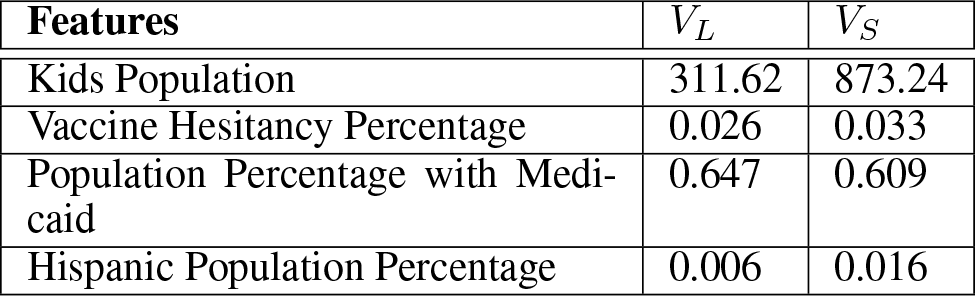
Average values of significant features in the set *V*_*L*_ (nodes with large errors) and *V*_*S*_ (nodes with small errors).

**Figure 4:**
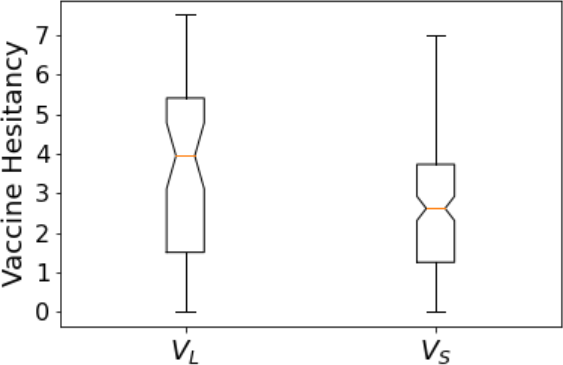
Vaccine hesitancy percentage in two sets, *V*_*L*_ and *V*_*S*_.

### 4.8 Forecasting Performance

We test the VH-GNN as a vaccine hesitancy forecasting tool. For this purpose, we train the VH-GNN until the *T* − 1 time step, then we use VH-GNN to forecast vaccine hesitancy percentages for all zip codes at time *T*. Figure 5 shows the forecast vaccine hesitancy percentage. The mean RMSE and MAE value for this case is 0.1602 and 0.1115.

**Figure 5:**
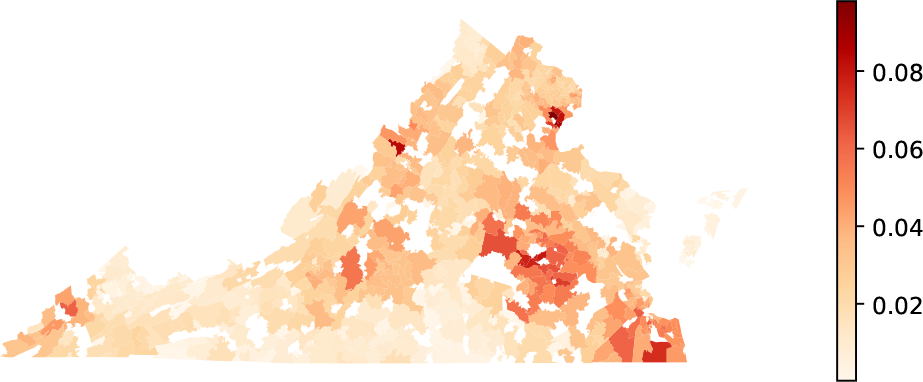
Vaccine hesitancy forecast for all the zip codes of Virginia in the first half of 2019. Here, colors represent vaccine hesitancy percentages.

### 4.9 Evaluation Metrics to Understand Spatial Structure

Understanding spatial structure is essential for the VaccHesitancy problem. We evaluate the performance of the VH-GNN in capturing spatial structure by using two following clustering measures:

- **Moran’s-I:** It is the measure of global spatial autocorrelation. This value ranges from −1 to 1, with 0 indicating no autocorrelation; −1 indicating perfect clustering with dissimilar values, such as clustering of high vaccine hesitancy location with the low vaccine hesitancy location; and 1 indicating perfect clustering with similar values, such as clustering of high vaccine hesitancy locations with high vaccine hesitancy locations. The equation for the Moran’s-I for a time *t* is

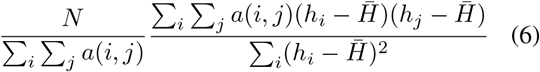
- **Isolation Index:** It indicates the level of segregation within a specific group or cluster compared to the larger population, with values ranging from 0 (no segregation) to 1 (full segregation). The equation for the isolation index for a time *t* is

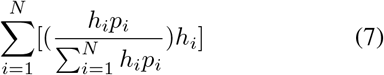

Our predicted value from the VaccHesitancy produces a Moran’s I value of 0.8488 for the year 2019, and the actual true value results in a value of 0.4580. The predicted value and the true value both finds that individuals exhibiting higher levels of vaccine hesitancy are more likely to be situated closely in *G*_*m*_.

The calculated isolation index from the predicted value and the true value is 0.0466 and 0.0480 for the year 2019, both indicates almost no segregation.

## 5 Conclusions

The VH-GNN framework is able to predict the spatiotemporal aspects of vaccine hesitancy with a combined GNN and RNN structure. Our method crucially uses a very large all payers insurance dataset, and a detailed activity-based synthetic contact network. Our method outperforms several baseline methods in predicting vaccine hesitancy at a zip code level, in terms of the RMSE and MAE evaluation metrics.

We also demonstrate the model’s effectiveness at the node-level data, highlighting the challenges in learning vaccine hesitancy for smaller populations. Although GRU is well-known to handle sequential data, they are computationally expensive and require a lot of data to train. We find that GRU or neural network alone cannot predict vaccine hesitancy at a zip code level. However, a combination of GNN and GRU can learn the spatial and temporal aspects of vaccine hesitancy and can predict patient refusal at a higher spatial resolution.

## Data Availability

All data produced in the present work are contained in the manuscript

